## Supplement for "Evidence of Leaky Protection Following COVID-19 Vaccination and SARS-CoV-2 Infection in an Incarcerated Population"

**DOC COVID-19 Testing**

To mitigate and track the spread of SARS-CoV-2, the Connecticut DOC conducts testing among residents for the following reasons: mass screening, symptom presence, contact tracing, movements (intakes and transfer), court hearings, and employment related reasons. The specifics for each reason are provided below:

- Mass screening (RT-PCR): Mass screening was conducted using RT-PCRs among a random sample of 10% of the resident population biweekly and was voluntary.
- Symptomatic Testing (Rapid Antigen Test): Residents who presented with COVID-19 symptoms without alternative reasons for the symptoms were tested with a rapid antigen test on the day the symptoms were reported.
- Contact tracing (Rapid Antigen Test): Contact tracing was performed among (1) residents with an infected cellmate, regardless of if contact was reported, or (2) residents with an infected cellblock- or facility-mate if close contact was reported by the infected resident. In accordance with the CDC, close contact was defined as being within six feet of an infected person for at least 15 minutes within a 24-hour period during the 0-2 days prior to symptom onset (or test confirmation in the absence of symptoms). The testing protocol stated that identified contacts were tested on the fifth day after close contact with the infected resident, but variation in the exact day of testing was likely, especially when infection burden was high. At the time of this analysis, testing capacity allowed for symptomatic and asymptomatic close contact evaluation and no prioritization was required.^1^
- Intake (Rapid Antigen Test): Residents were tested on day of intake using a rapid antigen test, held in quarantine for 10 days and tested on day 10 using a rapid antigen test.
- Inter-facility transfers: Residents transferred between facilities were tested using a rapid antigen test prior to transfer.
- Court hearings (Rapid Antigen Test): Residents were tested prior to court hearings using a rapid antigen test.
- Employment (Rapid Antigen Test): Some residents work within facilities or have community facing jobs that require regular SARS-CoV-2 testing. The specifics about the frequency of this testing varies by position.

**Housing Structure of Connecticut Correctional Facilities**

**Supplement Figure 1: *Depiction of within correctional facility housing structure***

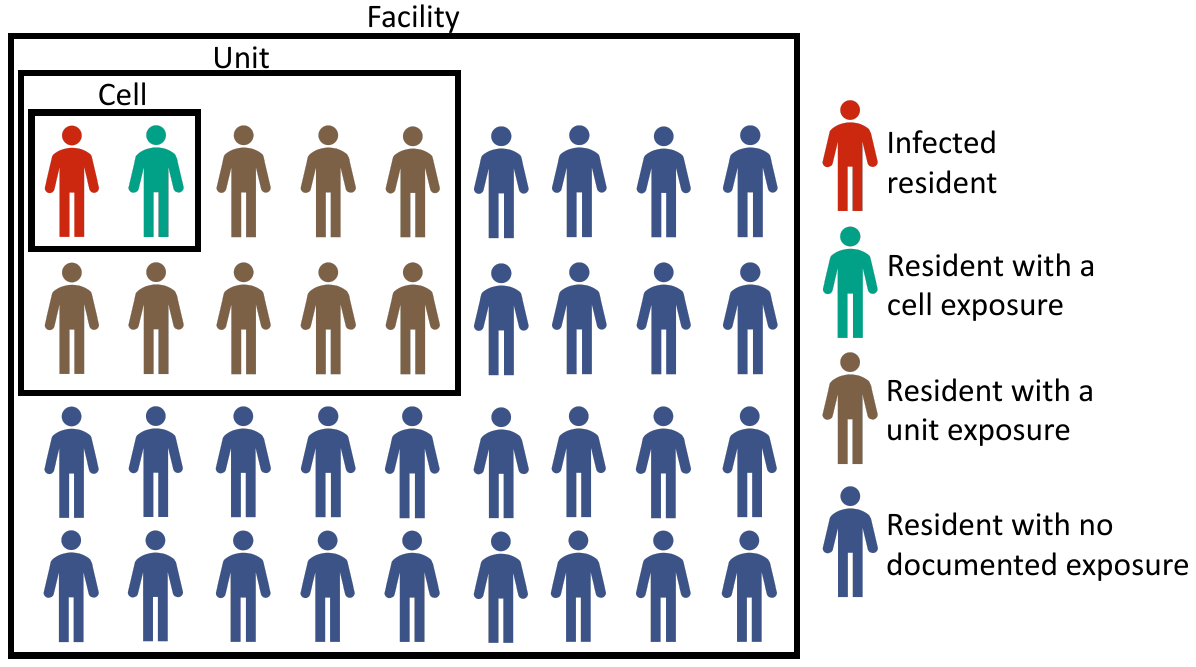

Legend: Illustration of how residents are organized within correctional facilities and how we defined residents as having a cell exposure event, cellblock exposure event, or an event without documented exposures. People who resided in cells between June 15, 2021, and May 10, 2022, were identified and classified as having a cell exposure event (green) if their cellmate tested positive for SARS-CoV-2 in the absence of a cell exposure event in the prior 14 days, having a cellblock exposure event (brown) if a resident of their cellblock but not their cell tested positive for SARS-CoV-2 in the absence of a cell or cellblock exposure event in the prior 14 days, and having no documented exposure (navy) if no one in their cellblock or cell tested positive for SARS-CoV-2 on a given day or in the prior 14 days. The human vector used in the generation of this figure was taken from Microsoft PowerPoint for Mac version 16.63.

| **Supplemental Table 1: Characteristics of Residents with and without Documented Exposure to SARS-CoV-2 Infected Residents by Gender** | | | | | | | | | | |
| --- | --- | --- | --- | --- | --- | --- | --- | --- | --- | --- |
|  | Delta Predominant Period (June 15, 2021 – Dec. 12, 2022) | | | Omicron Predominant Period (Dec. 13, 2022 - May 10, 2022) | | | | | | |
| Characteristics | Cell exposure events | Cellblock exposure events | Events without documented exposures | Cell exposure events | | | Cellblock exposure events | | | Events without documented exposures |
| MALE | | | | | | | | | | |
|  | (N=226) | (N=5233) | (N=15549) | (N=591) | | (N=5106) | | | (N=12001) | |
| Age (Median [Qr 1-3]) | 34 [25, 43] | 36 [29, 46] | 36 [29, 46] | 37 [29, 47] | | 36 [27, 45] | | | 37 [29, 46] | |
| Gender Female (N [%]) | 226 (100%) | 5233 (100%) | 15549 (100%) | 591 (100%) | | 5106 (100%) | | | 12001 (100%) | |
| Race (N [%]) |  |  |  |  | |  | | |  | |
| Non-Hispanic Black | 118 (52.2%) | 2378 (45.4%) | 7331 (47.1%) | 279 (47.2%) | | 2405 (47.1%) | | | 5633 (46.9%) | |
| Non-Hispanic White | 50 (22.1%) | 1294 (24.7%) | 3635 (23.4%) | 126 (21.3%) | | 1159 (22.7%) | | | 2739 (22.8%) | |
| Other (Hispanic, American Indian,  & Asian) | 58 (25.7%) | 1561 (29.8%) | 4583 (29.5%) | 186 (31.5%) | | 1542 (30.2%) | | | 3629 (30.2%) | |
| Duration of Incarceration (Median [QR 1-3])^a^ | 181 [178, 181] | 181 [181, 181] | 181 [181, 181] | 149 [149, 149] | | 149 [149, 149] | | | 149 [149, 149] | |
| Cell Size (Median [QR 1-3])^b^ | 2 [2, 2] | 2 [2, 2] | 2 [2, 2] | 2 [2, 2] | | 2 [2, 2] | | | 2 [2, 2] | |
| Cellblock Size (Median [QR 1-3])^b^ | 86 [55, 138] | 128 [69, 155] | 91 [67, 147] | 88 [62, 133] | | 93 [70, 152] | | | 92 [68, 126] | |
| History of Prior Infection (N [%])^c^ | 80 (35.4%) | 2120 (40.5%) | 6209 (39.9%) | 232 (39.3%) | | 2410 (47.2%) | | | 5975 (49.8%) | |
| History of Vaccination (N [%])^c^ | 95 (42.0%) | 2839 (54.3%) | 8367 (53.8%) | 334 (56.5%) | | 2998 (58.7%) | | | 7033 (58.6%) | |
| History of Hybrid Immunity (N [%])^c^ | 41 (18.1%) | 1335 (25.5%) | 4047 (26.0%) | 157 (26.6%) | | 1595 (31.2%) | | | 3857 (32.1%) | |
| Follow-up Time (Median [Qr 1-3]) | 14 [14, 14] | 14 [14, 14] | 14 [4, 14] | 14 [14, 14] | | 14 [14, 14] | | | 10 [3, 14] | |
| FEMALE | | | | | | | | | | |
|  | (N=38) | (N=383) | (N=1475) | (N=111) | (N=874) | | | (N=1463) | | |
| Age (Median [Qr 1-3]) | 37 [33, 42] | 36 [30, 42] | 35 [31, 42] | 36 [31, 43] | 35 [30, 42] | | | 35 [30, 42] | | |
| Gender Female (N [%]) | 38 (100%) | 383 (100%) | 1475 (100%) | 111 (100%) | 874 (100%) | | | 1463 (100%) | | |
| Race (N [%]) |  |  |  |  |  | | |  | | |
| Non-Hispanic Black | 8 (21.1%) | 106 (27.7%) | 405 (27.5%) | 39 (35.1%) | 239 (27.3%) | | | 419 (28.6%) | | |
| Non-Hispanic White | 19 (50.0%) | 188 (49.1%) | 719 (48.7%) | 48 (43.2%) | 424 (48.5%) | | | 686 (46.9%) | | |
| Other (Hispanic, American Indian,  & Asian) | 11 (28.9%) | 89 (23.2%) | 351 (23.8%) | 24 (21.6%) | 211 (24.1%) | | | 358 (24.5%) | | |
| Duration of Incarceration (Median [QR 1-3])^a^ | 97.5 [74.3, 181] | 144 [73.5, 181] | 181 [98.0, 181] | 149 [112, 149] | 149 [114, 149] | | | 149 [109, 149] | | |
| Cell Size (Median [QR 1-3])^b^ | 2 [2, 3] | 2 [2, 2] | 2 [2, 2] | 2 [2, 3] | 2 [2, 3] | | | 2. [2, 3] | | |
| Cellblock Size (Median [QR 1-3])^b^ | 69 [63, 77] | 77 [69, 87] | 65 [58, 73] | 65 [61, 71] | 69 [63, 77] | | | 69 [60, 77] | | |
| History of Prior Infection (N [%])^c^ | 5 (13.2%) | 60 (15.7%) | 313 (21.2%) | 22 (19.8%) | 196 (22.4%) | | | 354 (24.2%) | | |
| History of Vaccination (N [%])^c^ | 14 (36.8%) | 174 (45.4%) | 774 (52.5%) | 45 (40.5%) | 403 (46.1%) | | | 660 (45.1%) | | |
| History of Hybrid Immunity (N [%])^c^ | 4 (10.5%) | 44 (11.5%) | 242 (16.4%) | 11 (9.9%) | 134 (15.3%) | | | 238 (16.3%) | | |
| Follow-up Time (Median [Qr 1-3]) | 14 [14, 14] | 14 [14, 14] | 14 [4, 14] | 14 [13, 14] | 14 [12, 14] | | | 4 [1, 14] | | |
| ^a^ Duration of incarceration (in days) including the SARS-CoV-2 exposure event type | | | | | | | | | | |
| ^b^ Number of cellmates or unit-mates at the time of the exposure event that resulted in study inclusion | | | | | | | | | | |
| ^c^ History of prior infection defined as a record of prior infection (defined as prior infection defined as the record of a positive SARS-CoV-2 test (rapid antigen or RT-PCR) at least 90 days prior to follow-up start date); history of vaccination defined a receipt of at least one vaccine dose as of the start of follow-up; hybrid immunity defined as a recorded prior infection and vaccination as of the start of the study period | | | | | | | | | | |

| **Supplement Table 2: Difference in time since last prior infection or vaccine dose by facility event type^a^** | | | | | | | |
| --- | --- | --- | --- | --- | --- | --- | --- |
|  | Delta Predominant Period (June 15, 2021 - December 12, 2022) | | | | Omicron Predominant Period (December 123, 2022 - May 10, 2022) | | |
|  | Difference in Mean Days | 95% CI | Pvalue | Difference in Mean Days | | 95% CI | Pvalue |
| Time Since Last Prior Infection^b^ |  |  |  |  | |  | - |
| Events without documented exposures | - | - | - | - | | - | - |
| Cellblock exposure events | 3.9 | (-0.6, 8.4) | 0.087 | 6.38 | | (-0.7, 13.5) | 0.079 |
| Cell exposure events | 2.3 | (-16.8, 21.4) | 0.812 | 14.212 | | (-4.7, 33.1) | 0.141 |
| Time Since Last Vaccine Dose^b^ |  |  |  |  | |  |  |
| Events without documented exposures | - | - | - | - | | - | - |
| Cellblock exposure events | -0.6 | (-3.6, 2.3) | 0.671 | 1.1 | | (-3.5, 5.7) | 0.644 |
| Cell exposure events | 9.0 | (-4.3, 22.2) | 0.148 | -3.5 | | (-14.9, 8.0) | 0.553 |
| ^a^ Model adjusted for age, race, room size, date of event, and housing cellblock, hypothesis testing was performed using two-sided z-tests and no multiple comparison adjustment was performed | | | | | | | |
| ^b^ History of prior infection defined as a record of prior infection (defined as prior infection defined as the record of a positive SARS-CoV-2 test (rapid antigen or RT-PCR) at least 90 days prior to follow-up start date); history of vaccination defined a receipt of at least one vaccine dose as of the start of follow-up; hybrid immunity defined as a recorded prior infection and vaccination as of the start of the study period | | | | | | | |

| **Supplement Table 3: Unadjusted (primary analysis) estimates of the association between documented exposure and SARS-CoV-2 infection risk among residents of Connecticut Department of Correction facilities between June 15, 2021, and May 10, 2022** | | |
| --- | --- | --- |
| **Type of Facility Exposure** | **Hazard Ratio** | **95% Confidence Intervals** |
| Delta Period (June 15 to Dec. 12, 2021) | | |
| No Exposure | - | - |
| Cellblock Exposure | 4.58 | (3.68-5.70) |
| Cell Exposures | 24.13 | (17.48-33.32) |
| Omicron Period (Dec. 13, 2021, to May 10, 2022) | | |
| No Exposure | - | - |
| Cellblock Exposure | 4.79 | (4.02-5.71) |
| Cell Exposures | 8.87 | (6.97-11.30) |

**Outcome of Symptomatic Infection:**

**High exposure setting overcomes the protection afforded by infection, vaccination, and hybrid immunity against symptomatic infection**

As a secondary analyses, we estimated the effect of prior infection, vaccination, and hybrid immunity against symptomatic infection. Because symptom data was only available for rapid antigen tests, we defined a symptomatic infection as a positive rapid antigen test collected from a symptomatic resident. We used the same models for these analyses as we did for the infection outcome analyses.

**Supplement Figure 2: *Association between documented SARS-CoV-2 exposure and symptomatic SARS-CoV-2 infection risk among residents of Connecticut Department of Correction Facilities between June 15, 2021, and May 10, 2022***

Legend: Forest plot depicting the association between documented close exposure to a SARS-CoV-2 infected resident and the risk of subsequent symptomatic SARS-CoV-2 infection. Residents were classified as having a cell exposure event (green) on the day their cellmate tested positive, having a cellblock exposure event (brown) the day a resident of their cellblock but not cell tested positive, and having an event without documented exposures if no one in their cellblock tested positive on a given day. Cell exposure events that occurred within 14 days following a prior cell exposure event were excluded. Cellblock exposure events and events without documented exposures that occurred in the 14 days following a cellblock or cell exposure event were excluded. Facility exposures were stratified by periods of variant predominance (Delta [A]: June 15, 2021 – December 12, 2021; Omicron [B]: December 13, 2021 – May 10, 2022). The associations were estimated using a Cox Proportional Hazard Model stratified by facility and with robust standard errors. The model was adjusted for age, calendar date, race, room and cellblock size, vaccination, and prior infection status of the susceptible person. Boxes indicate estimated hazard ratio (HR) point values and whiskers indicate 95% confidence intervals (Delta Period: n = 22,904 facility events; Omicron Period: n = 20,146 facility events).

| **Supplement Table 4: Unadjusted estimates of the association between documented exposure and symptomatic SARS-CoV-2 infection risk among residents of Connecticut Department of Correction facilities between June 15, 2021, and May 10, 2022** | | |
| --- | --- | --- |
| **Type of Facility Exposure** | **Hazard Ratio** | **95% Confidence Intervals** |
| Delta Period (June 15 to Dec. 12, 2021) | | |
| No Exposure | - | - |
| Cellblock Exposure | 3.61 | (2.61-5.00) |
| Cell Exposures | 18.17 | (11.05-29.87) |
| Omicron Period (Dec. 13, 2021 to May 10, 2022) | | |
| No Exposure |  |  |
| Cellblock Exposure | 4.44 | (3.41-5.78) |
| Cell Exposures | 10.21 | (7.25-14.38) |

**Supplement Figure 3: *Effectiveness of prior infection, vaccination, and hybrid immunity on symptomatic SARS-CoV-2 infection among residents of Connecticut Department of Correction facilities between June 15, 2021, and May 10, 2022, by documented exposure status***

Legend: Forest plot depicting the association between prior infection, vaccination, and hybrid immunity and the risk of symptomatic SARS-CoV-2 infection by facility exposure type. Residents were classified as having a cell exposure event (green) the day their cellmate tested positive, having a cellblock exposure event (brown) the day a resident of their cellblock but not cell tested positive, and having an event without documented exposure (navy) if no one in their cellblock tested positive. Cell exposure events that occurred within 14 days following a prior cell exposure event were excluded. Cellblock exposure events and events without documented exposure that occurred in the 14 days following a cellblock or cell exposure event were excluded. Associations were examined using Cox Proportional Hazard Models stratified by cellblock with robust standard errors. Each model was adjusted for age, date of exposure, race, room size, and model (a) was adjusted for vaccination status and model (b) was adjusted for prior infection status. Model (c) was limited to residents with hybrid immunity or residents without a record of prior infection or vaccination. Prior infection was defined as a recorded positive SARS-CoV-2 test ≥90 days before the event and vaccination was defined as the receipt of ≥1 dose before the event. Hybrid immunity was defined as a record of both a prior infection and ≥1 vaccine dose. Boxes indicate estimated hazard ratio (HR) point values and whiskers indicate 95% confidence intervals (Delta: n = 17,024 no exposure events, 5,616 cellblock exposure events, 264 cell exposure events; Omicron: n = 13,464 no exposure events, 5,980 cellblock exposure events, 702 cell exposure events). Ratio of HRs refer to the p-value comparing the HR following cellblock or cell exposure events to the HR following events without documented exposures, estimated using two-sided z-tests. No multiple testing adjustment was performed.

| **Supplement Table 5: *Unadjusted estimates of the effectiveness of prior infection, vaccination, and hybrid immunity on symptomatic SARS-CoV-2 infection among residents of Connecticut Department of Correction facilities between June 15, 2021, and May 10, 2022, by documented exposure status*** | | | | | | |
| --- | --- | --- | --- | --- | --- | --- |
|  | **Delta Period (June 15 to Dec. 12, 2021)** | | | **Omicron Period (Dec. 13, 2021 - May 10, 2022)** | | |
| **Type of Facility Exposure** | **Hazard Ratio** | **95% Confidence Intervals** | **Ratio of HR (Pvalue)^a^** | **Hazard Ratio** | **95% Confidence Intervals** | **Ratio of HR (Pvalue)^a^** |
| Prior SARS-CoV-2 Infection^b^ | | | | | | |
| No Exposure | 0.15 | (0.06-0.37) | - | 0.33 | (0.20-0.56) | - |
| Cellblock Exposure | 0.26 | (0.14-0.47) | 0.304 | 0.68 | (0.51-0.91) | 0.020 |
| Cell Exposures | 0.33 | (0.10-1.13) | 0.289 | 0.91 | (0.53-1.57) | 0.009 |
| Prior Vaccination^c^ | | | | | | |
| No Exposure | 0.21 | (0.11-0.40) | - | 0.31 | (0.19-0.50) | - |
| Cellblock Exposure | 0.19 | (0.12-0.33) | 0.841 | 0.69 | (0.53-0.91) | 0.007 |
| Cell Exposures | 0.43 | (0.16-1.17) | 0.243 | 0.82 | (0.49-1.38) | 0.005 |
| Hybrid Immunity^d^ | | | | | | |
| No Exposure | 0.03 | (0.00-0.21) | - | 0.13 | (0.06-0.29) | - |
| Cellblock Exposure | 0.06 | (0.02-0.55) | 0.549 | 0.52 | (0.35-0.75) | 0.030 |
| Cell Exposures | - | - | - | 0.80 | (0.40-1.59) | 0.001 |
| ^a^ Hypothesis testing was performed using two-sided z-tests and no multiple comparison adjustment was performed | | | | | | |
| ^b^ Prior infections were defined as a recorded positive SARS-CoV-2 test at least 90 days prior to the event and vaccination was defined as the receipt of at least one dose prior to the event. | | | | | | |
| ^c^ Residents were classified as being vaccinated if they had received at least one vaccine dose. | | | | | | |
| ^d^ Hybrid immunity was defined as a record of both a prior infection and at least one vaccine dose. No infections among cell exposure group. | | | | | | |

**Proportion of residents with cell exposure events, cellblock exposure events and events without documented exposure by testing reason**

**Supplement Figure 4: Testing proportion by facility exposure and testing reason**

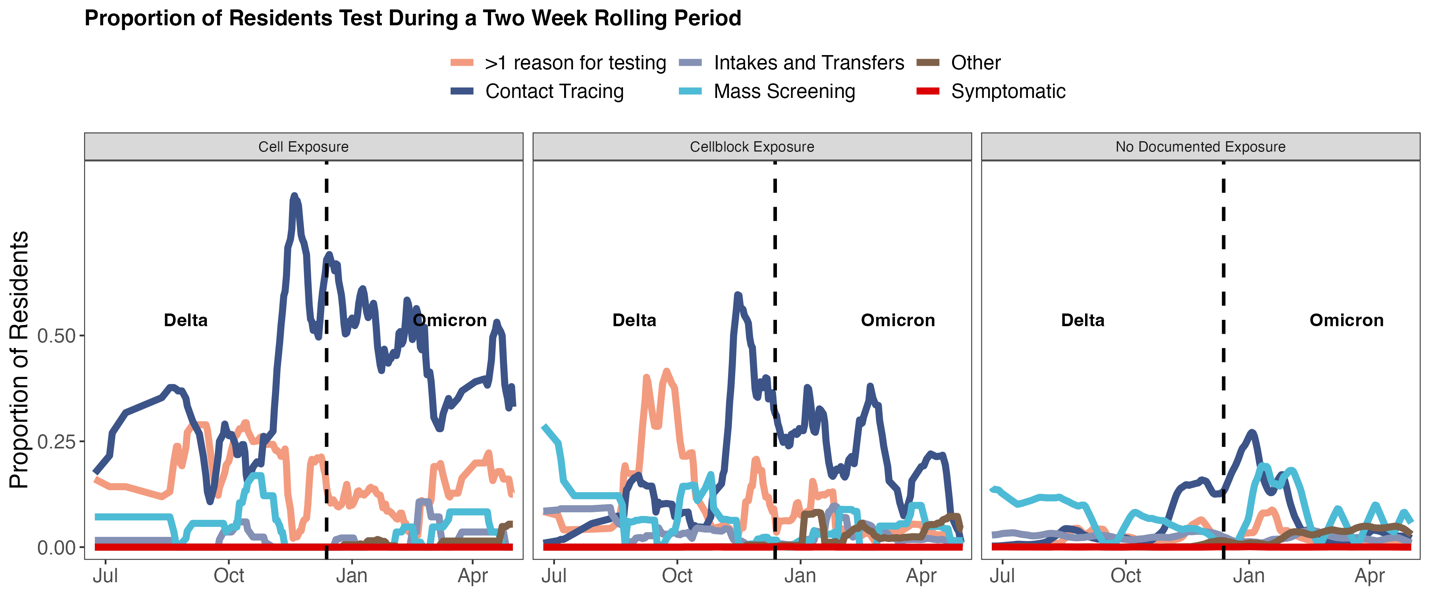

Legend: Proportion of residents tested over the study period by reason for testing, and facility exposure type. Residents were classified as having a cell exposure event the day their cellmate tested positive, having a cellblock exposure event the day a resident of their cellblock but not cell tested positive, and having an event without documented exposure if no one in their cellblock tested positive. Cell exposure events that occurred within 14 days following a prior cell exposure event were excluded. Cellblock exposure events and events without documented exposure that occurred in the 14 days following a cellblock or cell exposure event were excluded. Residents were tested for the following reasons: mass screening (light blue), contact tracing in the absence of recorded symptoms (navy), intake/transfer testing in the absence of recorded symptoms (grey/blue), other testing in the absence of recorded symptoms (brown), testing in the presence of recorded symptoms (symptoms data not available for mass screening [PCR] testing; red), and tested for more than one reason on the same day (pink).

**Comparison in the effect of prior infection and vaccination against SARS-CoV-2 infection between events with and without documented exposures**

**Supplement Table 6. *Ratio of hazard ratios following cellblock and cell exposure events to events without documented exposures^a^***

| Prior SARS-CoV-2 Infection^b^ | | | | | | | | | |
| --- | --- | --- | --- | --- | --- | --- | --- | --- | --- |
|  |  | Delta Predominant Period | | | | Omicron Predominant Period | | | |
|  |  | Ratio of HR | 95% CI | P-Value | Ratio of HR | | 95% CI | | P-Value |
|  | No Exposure^c^ | Ref | Ref | Ref | Ref | | Ref | | Ref |
|  | Cellblock Exposure^c^ | 1.55 | (0.77-3.12) | 0.216 | 1.10 | | (0.65- 1.84) | | 0.727 |
|  | Cell Exposure^c^ | 2.80 | (1.11-7.09) | 0.029 | 2.33 | | (1.07-5.08) | | 0.033 |
|  | Prior Vaccination^d^ | | | | | | | | |
|  |  | Delta Predominant Period | | | | Omicron Predominant Period | | | |
|  |  | Ratio of HR | 95% CI | P-Value | Ratio of HR | | 95% CI | P-Value | |
|  | No Exposure^c^ | Ref | Ref | Ref | Ref | | Ref | Ref | |
|  | Cellblock Exposure^c^ | 1.67 | (1.08-2.57) | 0.019 | 1.20 | | (0.84-1.71) | 0.313 | |
|  | Cell Exposure^c^ | 2.44 | (1.39-4.26) | 0.002 | 1.68 | | (1.02-2.76) | 0.041 | |
|  | Hybrid Immunity^e^ | | | | | | | | |
|  |  | Delta Predominant Period | | | | Omicron Predominant Period | | | |
|  |  | Ratio of HR | 95% CI | P-Value | Ratio of HR | | 95% CI | | P-Value |
|  | No Exposure^c^ | Ref | Ref | Ref | Ref | | Ref | | Ref |
|  | Cellblock Exposure^c^ | 2.01 | (1.24-29.72) | 0.203 | 1.72 | | (0.99-2.96) | | 0.053 |
|  | Cell Exposure^c^ | 6.08 | (1.39-4.26) | 0.026 | 3.33 | | (1.64-6.78) | | 0.001 |
|  | ^a^ Hypothesis testing was performed using two-sided z-tests and no multiple comparison adjustment was performed | | | | | | | | |
|  | ^b^ Prior infections were defined as a recorded positive SARS-COV-2 test at least 90 days prior to the event; model adjusted for age, date of exposure, race, room size, and vaccination status | | | | | | | | |
|  | ^c^ Residents were classified as having a cell exposure event on the day their cellmate tested positive, having a cellblock exposure event the day a resident of their cellblock but not cell tested positive, and having an event without documented exposures if no one in their cellblock tested positive on a given day. Cell exposure events that occurred within 14 days following a prior cell exposure event were excluded. Cellblock exposure events and events without documented exposures that occurred in the 14 days following a cellblock or cell exposure event were excluded. | | | | | | | | |
|  | ^d^ Vaccination was defined as the receipt of at least one vaccine dose prior to the event; model adjusted for age, date of exposure, race, room size, and prior infection status | | | | | | | | |
|  | ^e^ Hybrid immunity defined as having both a prior infection and vaccine dose record; model adjusted for age, date of exposure, race, and room size. Limited to residents with hybrid immunity or no history of prior infection or vaccination | | | | | | | | |

| **Supplement Table 7: *Unadjusted (primary analysis) estimates of the effectiveness of prior infection, vaccination, and hybrid immunity on SARS-CoV-2 infection among residents of Connecticut Department of Correction facilities between June 15, 2021, and May 10, 2022, by documented exposure status*** | | | | | | |
| --- | --- | --- | --- | --- | --- | --- |
|  | **Delta Period (June 15 to Dec. 12, 2021)** | | | **Omicron Period (Dec. 13, 2021 - May 10, 2022)** | | |
| **Type of Facility Exposure** | **Hazard Ratio** | **95% Confidence Intervals** | **Ratio of HR (Pvalue)^a^** | **Hazard Ratio** | **95% Confidence Intervals** | **Ratio of HR (Pvalue)^a^** |
| Prior SARS-CoV-2 Infection^b^ | | | | | | |
| No Exposure | 0.16 | (0.09-0.30) | - | 0.31 | (0.22-0.45) | - |
| Cellblock Exposure | 0.26 | (0.18-0.38) | 0.170 | 0.57 | (0.47-0.69) | 0.004 |
| Cell Exposures | 0.57 | (0.30-1.09) | 0.005 | 0.84 | (0.57-1.26) | <0.001 |
| Prior Vaccination^c^ | | | | | | |
| No Exposure | 0.28 | (0.18-0.42) | - | 0.51 | (0.38-0.70) | - |
| Cellblock Exposure | 0.32 | (0.24-0.43) | 0.575 | 0.71 | (0.60-0.85) | 0.069 |
| Cell Exposures | 0.64 | (0.36-1.14) | 0.027 | 1.05 | (0.72-1.54) | 0.004 |
| Hybrid Immunity^d^ | | | | | | |
| No Exposure | 0.06 | (0.02-0.17) | - | 0.21 | (0.13-0.34) | - |
| Cellblock Exposure | 0.10 | (0.06-0.19) | 0.380 | 0.45 | (0.35-0.58) | 0.006 |
| Cell Exposures | 0.33 | (0.12-0.94) | 0.023 | 0.90 | (0.53-1.50) | <0.001 |
| ^a^ Hypothesis testing was performed using two-sided z-tests and no multiple comparison adjustment was performed | | | | | | |
| ^b^ Prior infections were defined as a recorded positive SARS-CoV-2 test at least 90 days prior to the event and vaccination was defined as the receipt of at least one dose prior to the event. | | | | | | |
| ^c^ Residents were classified as being vaccinated if they had received at least one vaccine dose. | | | | | | |
| ^d^ Hybrid immunity was defined as a record of both a prior infection and at least one vaccine dose. | | | | | | |

| **Supplement Table 8: *Unadjusted (primary analysis) estimates of the effectiveness of prior infection and vaccination status of index cases on SARS-CoV-2 transmissibility among of residents of Connecticut Department of Correction facility between June 15, 2021, and May 10, 2022, by documented SARS-CoV-2 exposure status*** | | | | |
| --- | --- | --- | --- | --- |
|  | **Delta Period (June 15 to Dec. 12, 2021)** | | **Omicron Period (Dec. 13, 2021 - May 10, 2022)** | |
| **Type of Facility Exposure** | **Hazard Ratio** | **95% Confidence Intervals** | **Hazard Ratio** | **95% Confidence Intervals** |
| Prior SARS-CoV-2 Infection^a^ | | | | |
| Cellblock Exposure | 0.55 | (0.36-0.85) | 0.85 | (0.68-1.06) |
| Cell Exposures | 0.98 | (0.48-2.01) | 1.36 | (0.94-1.97) |
| Prior Vaccination^b^ | | | | |
| Cellblock Exposure | 0.56 | (0.40-0.79) | 0.96 | (0.80-1.15) |
| Cell Exposures | 1.06 | (0.58-1.92) | 1.19 | (0.83-1.70) |
| ^a^ Prior infections were defined as a recorded positive SARS-CoV-2 test at least 90 days prior to the event and vaccination was defined as the receipt of at least one dose prior to the event. | | | | |
| ^b^ Residents were classified as being vaccinated if they had received at least one vaccine dose. | | | | |

**Sensitivity Analyses:**

***Restricted to people tested during 14-day follow-up***

In the primary analysis, we included residents regardless of if they were tested for SARS-CoV-2 during their 14 days following an event. Because symptomatic testing comprised a small proportion of testing (see previous figure and table), we anticipated the amount of bias introduced by including all residents regardless of testing status would be limited. However, as the frequency of testing differs between people with and without known exposures (Figure 1.C), there is a chance that we introduced bias into our analysis through this broad inclusion. Here, we restrict to events where residents were tested during follow-up.

**Supplement Figure 5: *Association between documented SARS-CoV-2 exposure and SARS-CoV-2 infection risk among residents of Connecticut Department of Correction Facilities between June 15, 2021, and May 10, 2022 (restricted to tested people)***

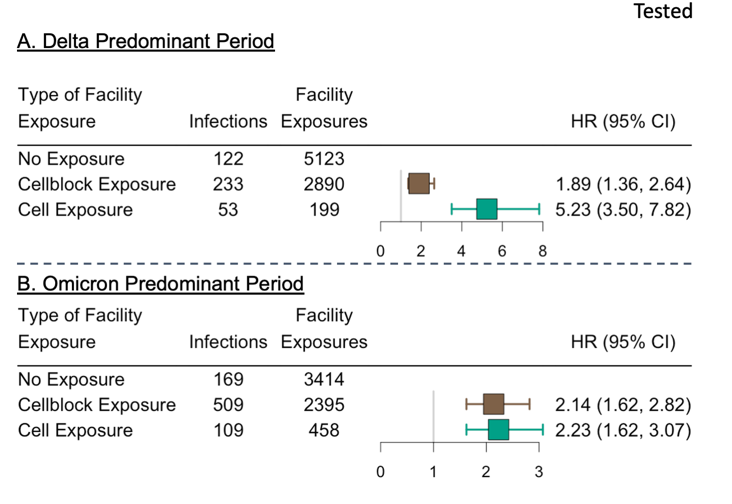
Legend: Forest plot depicting the association between documented close exposure to a SARS-CoV-2 infected resident and the risk of subsequent SARS-CoV-2 infection for residents tested during 14-day follow-up. Residents were classified as having a cell exposure event (green) on the day their cellmate tested positive, having a cellblock exposure event (brown) the day a resident of their cellblock but not cell tested positive, and having an event without documented exposures if no one in their cellblock tested positive on a given day. Cell exposure events that occurred within 14 days following a prior cell exposure event were excluded. Cellblock exposure events and events without documented exposures that occurred in the 14 days following a cellblock or cell exposure event were excluded. Facility exposures were stratified by periods of variant predominance (Delta [A]: June 15, 2021 – December 12, 2021; Omicron [B]: December 13, 2021 – May 10, 2022). The associations were estimated using a Cox Proportional Hazard Model stratified by facility and with robust standard errors. The model was adjusted for age, calendar date, race, room and cellblock size, vaccination, and prior infection status of the susceptible person. Boxes indicate estimated hazard ratio (HR) point values and whiskers indicate 95% confidence intervals (Delta Period: n = 8,212 facility events; Omicron Period: n = 6,267 facility events).

| **Supplement Table 9: *Unadjusted estimates of the association between documented exposure and SARS-CoV-2 infection risk among tested residents*** | | |
| --- | --- | --- |
| **Type of Facility Exposure** | **Hazard Ratio** | **95% Confidence Intervals** |
| Delta Period (June 15 to Dec. 12, 2021) | | |
| No Exposure | - | - |
| Cellblock Exposure | 2.17 | (1.74-2.71) |
| Cell Exposures | 7.94 | (5.75-10.96) |
| Omicron Period (Dec. 13, 2021, to May 10, 2022) | | |
| No Exposure | - | - |
| Cellblock Exposure | 2.74 | (2.30-3.27) |
| Cell Exposures | 2.9 | (2.28-3.70) |

**Supplement Figure 6: *Effect of prior infection, vaccination, and hybrid immunity on the susceptibility of residents of Connecticut Department of Correction Facilities to SARS-CoV-2 infection between June 15, 2021, and May 10, 2022, by documented SARS-CoV-2 exposure status*** ***(restricted to tested people)***

Legend: Forest plot depicting the association between prior infection, vaccination, and hybrid immunity and the risk of SARS-CoV-2 infection by facility exposure type for residents tested during 14-day follow-up. Residents were classified as having a cell exposure event (green) the day their cellmate tested positive, having a cellblock exposure event (brown) the day a resident of their cellblock but not cell tested positive, and having an event without documented exposure (navy) if no one in their cellblock tested positive. Cell exposure events that occurred within 14 days following a prior cell exposure event were excluded. Cellblock exposure events and events without documented exposure that occurred in the 14 days following a cellblock or cell exposure event were excluded. Associations were examined using Cox Proportional Hazard Models stratified by cellblock with robust standard errors. Each model was adjusted for age, date of exposure, race, room size, and model (a) was adjusted for vaccination status and model (b) was adjusted for prior infection status. Model (c) was limited to residents with hybrid immunity or residents without a record of prior infection or vaccination. Prior infection was defined as a recorded positive SARS-CoV-2 test ≥90 days before the event and vaccination was defined as the receipt of ≥1 dose before the event. Hybrid immunity was defined as a record of both a prior infection and ≥1 vaccine dose. Boxes indicate estimated hazard ratio (HR) point values and whiskers indicate 95% confidence intervals (Delta: n = 5,123 no exposure events, 2,890 cellblock exposure events, 199 cell exposure events; Omicron: n = 3,414 no exposure events, 2,395 cellblock exposure events, 458 cell exposure events). Ratio of HRs refer to the p-value comparing the HR following cellblock or cell exposure events to the HR following events without documented exposures, estimated using two-sided z-tests. No multiple testing adjustment was performed.

| **Supplement Table 10: *Unadjusted estimates of the effectiveness of prior infection, vaccination, and hybrid immunity on SARS-CoV-2 infection among tested residents*** | | | | | | |
| --- | --- | --- | --- | --- | --- | --- |
|  | **Delta Period (June 15 to Dec. 12, 2021)** | | | **Omicron Period (Dec. 13, 2021 - May 10, 2022)** | | |
| **Type of Facility Exposure** | **Hazard Ratio** | **95% Confidence Intervals** | **Ratio of HR (Pvalue)^a^** | **Hazard Ratio** | **95% Confidence Intervals** | **Ratio of HR (Pvalue)^a^** |
| Prior SARS-CoV-2 Infection^b^ | | | | | | |
| No Exposure | 0.16 | (0.09-0.31) | - | 0.36 | (0.25-0.52) | - |
| Cellblock Exposure | 0.26 | (0.18-0.38) | 0.207 | 0.55 | (0.45-0.66) | 0.048 |
| Cell Exposures | 0.41 | (0.22-0.79) | 0.043 | 0.59 | (0.40-0.88) | 0.073 |
| Prior Vaccination^c^ | | | | | | |
| No Exposure | 0.24 | (0.15-0.36) | - | 0.46 | (0.34-0.64) | - |
| Cellblock Exposure | 0.28 | (0.21-0.38) | 0.481 | 0.56 | (0.47-0.67) | 0.322 |
| Cell Exposures | 0.51 | (0.29-0.91) | 0.032 | 0.75 | (0.52-1.11) | 0.051 |
| Hybrid Immunity^d^ | | | | | | |
| No Exposure | 0.05 | (0.02-0.15) | - | 0.22 | (0.14-0.35) | - |
| Cellblock Exposure | 0.09 | (0.05-0.17) | 0.385 | 0.35 | (0.27-0.45) | 0.086 |
| Cell Exposures | 0.21 | (0.07-0.61) | 0.067 | 0.50 | (0.30-0.83) | 0.021 |
| ^a^ Hypothesis testing was performed using two-sided z-tests and no multiple comparison adjustment was performed | | | | | | |
| ^b^ Prior infections were defined as a recorded positive SARS-CoV-2 test at least 90 days prior to the event and vaccination was defined as the receipt of at least one dose prior to the event. | | | | | | |
| ^c^ Residents were classified as being vaccinated if they had received at least one vaccine dose. | | | | | | |
| ^d^ Hybrid immunity was defined as a record of both a prior infection and at least one vaccine dose. No infections among cell exposure group. | | | | | | |

***Perform the analyses among residents with a recorded reason for testing other than symptomatic***

In the prior sensitivity analysis, we limited to residents tested during follow-up. While this reduces the risk of testing related bias, residual bias may be present due to differences in testing reason. Of specific concern was symptomatic testing. Because testing for symptomatic reasons may be associated with prior infection or vaccination statuses and is associated with the levels of protection offered by prior infection and vaccination, we wanted to examine if this could be driving our findings suggesting leaky vaccines. For this reason, we conducted an analysis restricted to residents with tests in the 14 days following inclusion performed for reasons other than symptomatic testing. Note, for this analysis, we did not exclude residents tested for reasons other than symptomatic testing, even if symptoms were recorded.

**Supplement Figure 7: *Association between documented SARS-CoV-2 exposure and SARS-CoV-2 infection risk among residents of Connecticut Department of Correction Facilities between June 15, 2021, and May 10, 2022 (tested for reasons other than symptoms)***

Legend: Forest plot depicting the association between documented close exposure to a SARS-CoV-2 infected resident and the risk of subsequent SARS-CoV-2 infection for residents tested for reasons other than symptoms during 14-day follow-up. Residents were classified as having a cell exposure event (green) on the day their cellmate tested positive, having a cellblock exposure event (brown) the day a resident of their cellblock but not cell tested positive, and having an event without documented exposures if no one in their cellblock tested positive on a given day. Cell exposure events that occurred within 14 days following a prior cell exposure event were excluded. Cellblock exposure events and events without documented exposures that occurred in the 14 days following a cellblock or cell exposure event were excluded. Facility exposures were stratified by periods of variant predominance (Delta [A]: June 15, 2021 – December 12, 2021; Omicron [B]: December 13, 2021 – May 10, 2022). The associations were estimated using a Cox Proportional Hazard Model stratified by facility and with robust standard errors. The model was adjusted for age, calendar date, race, room and cellblock size, vaccination, and prior infection status of the susceptible person. Boxes indicate estimated hazard ratio (HR) point values and whiskers indicate 95% confidence intervals (Delta Period: n = 8,200 facility events; Omicron Period: n = 5,966 facility events).

| **Supplement Table 11: *Unadjusted estimates of the association between documented exposure and SARS-CoV-2 infection risk among residents tested for reasons other than symptoms*** | | |
| --- | --- | --- |
| **Type of Facility Exposure** | **Hazard Ratio** | **95% Confidence Intervals** |
| Delta Period (June 15 to Dec. 12, 2021) | | |
| No Exposure | - | - |
| Cellblock Exposure | 2.17 | (5.74-10.94) |
| Cell Exposures | 7.92 | (5.74-10.94) |
| Omicron Period (Dec. 13, 2021 to May 10, 2022) | | |
| No Exposure | - | - |
| Cellblock Exposure | 2.71 | (2.27-3.23) |
| Cell Exposures | 2.88 | (2.26-3.67) |

**Supplement Figure 8: *Effect of prior infection, vaccination, and hybrid immunity on the susceptibility of residents of Connecticut Department of Correction Facilities to SARS-CoV-2 infection between June 15, 2021, and May 10, 2022, by documented SARS-CoV-2 exposure status*** ***(tested for reasons other than symptoms)***

Legend: Forest plot depicting the association between prior infection, vaccination, and hybrid immunity and the risk of SARS-CoV-2 infection by facility exposure type for residents tested for reasons other than symptoms during 14-day follow-up. Residents were classified as having a cell exposure event (green) the day their cellmate tested positive, having a cellblock exposure event (brown) the day a resident of their cellblock but not cell tested positive, and having an event without documented exposure (navy) if no one in their cellblock tested positive. Cell exposure events that occurred within 14 days following a prior cell exposure event were excluded. Cellblock exposure events and events without documented exposure that occurred in the 14 days following a cellblock or cell exposure event were excluded. Associations were examined using Cox Proportional Hazard Models stratified by cellblock with robust standard errors. Each model was adjusted for age, date of exposure, race, room size, and model (a) was adjusted for vaccination status and model (b) was adjusted for prior infection status. Model (c) was limited to residents with hybrid immunity or residents without a record of prior infection or vaccination. Prior infection was defined as a recorded positive SARS-CoV-2 test ≥90 days before the event and vaccination was defined as the receipt of ≥1 dose before the event. Hybrid immunity was defined as a record of both a prior infection and ≥1 vaccine dose. Boxes indicate estimated hazard ratio (HR) point values and whiskers indicate 95% confidence intervals (Delta: n = 5,114 no exposure events, 2,887 cellblock exposure events, 199 cell exposure events; Omicron: n = 3,319 no exposure events, 2,195 cellblock exposure events, 452 cell exposure events). Ratio of HRs refer to the p-value comparing the HR following cellblock or cell exposure events to the HR following events without documented exposures, estimated using two-sided z-tests. No multiple testing adjustment was performed.

| **Supplement Table 12: *Unadjusted estimates of the effectiveness of prior infection, vaccination, and hybrid immunity on SARS-CoV-2 infection among residents tested for reasons other than symptoms*** | | | | | | | | | | | |
| --- | --- | --- | --- | --- | --- | --- | --- | --- | --- | --- | --- |
|  | **Delta Period (June 15 to Dec. 12, 2021)** | | | | | | **Omicron Period (Dec. 13, 2021 - May 10, 2022)** | | | | |
| **Type of Facility Exposure** | **Hazard Ratio** | **95% Confidence Intervals** | **Ratio of HR (Pvalue)^a^** | | **Hazard Ratio** | | | **95% Confidence Intervals** | | **Ratio of HR (Pvalue)^a^** | |
| Prior SARS-CoV-2 Infection^b^ | | | | | | | | | | | |
| No Exposure | 0.16 | (0.09-0.30) | | - | | 0.36 | | | (0.25-0.52) | | - |
| Cellblock Exposure | 0.26 | (0.18-0.38) | | 0.206 | | 0.54 | | | (0.45-0.66) | | 0.051 |
| Cell Exposures | 0.41 | (0.22-0.79) | | 0.042 | | 0.57 | | | (0.38-0.86) | | 0.095 |
| Prior Vaccination^c^ | | | | | | | | | | | |
| No Exposure | 0.23 | (0.16-0.36) | | - | | 0.47 | | | (0.34-0.64) | | - |
| Cellblock Exposure | 0.28 | (0.21-0.38) | | 0.481 | | 0.55 | | | (0.46-0.66) | | 0.354 |
| Cell Exposures | 0.51 | (0.29-0.91) | | 0.032 | | 0.78 | | | (0.53-1.14) | | 0.040 |
| Hybrid Immunity^d^ | | | | | | | | | | | |
| No Exposure | 0.05 | (0.02-0.15) | | - | | 0.22 | | | (0.14-0.35) | | - |
| Cellblock Exposure | 0.09 | (0.05-0.17) | | 0.384 | | 0.34 | | | (0.27-0.44) | | 0.094 |
| Cell Exposures | 0.21 | (0.07-0.61) | | 0.066 | | 0.49 | | | (0.30-0.83) | | 0.021 |
| ^a^ Hypothesis testing was performed using two-sided z-tests and no multiple comparison adjustment was performed | | | | | | | | | | | |
| ^b^ Prior infections were defined as a recorded positive SARS-CoV-2 test at least 90 days prior to the event and vaccination was defined as the receipt of at least one dose prior to the event. | | | | | | | | | | | |
| ^c^ Residents were classified as being vaccinated if they had received at least one vaccine dose. | | | | | | | | | | | |
| ^d^ Hybrid immunity was defined as a record of both a prior infection and at least one vaccine dose. No infections among cell exposure group. | | | | | | | | | | | |

***Restricted to people incarcerated since the beginning of the study (June 15, 2021)***

Because we did not have access to community infection records, residents who had a SARS-CoV-2 infection prior to incarceration may have been misclassified as not having a prior infection in our analysis. To examine the impact of these missing data, we conducted a sensitivity analysis restricted to people incarcerated since the beginning of the study (June 15, 2021). While we were unable to restrict the study population to people incarcerated since the beginning of the pandemic due to data limitations, infection induced seroprevalence estimates from Connecticut suggest that only between 4.7% and 19.7% of the state population had a SARS-CoV-2 infection prior to July 2021.^2^

**Supplement Figure 9: *Association between documented SARS-CoV-2 exposure and SARS-CoV-2 infection risk among residents of Connecticut Department of Correction Facilities between June 15, 2021, and May 10, 2022 (restricted to people incarcerated since beginning of study)***

Legend: Forest plot depicting the association between documented close exposure to a SARS-CoV-2 infected resident and the risk of subsequent SARS-CoV-2 infection for residents incarcerated since the beginning of the study (June 15, 2021). Residents were classified as having a cell exposure event (green) on the day their cellmate tested positive, having a cellblock exposure event (brown) the day a resident of their cellblock but not cell tested positive, and having an event without documented exposures if no one in their cellblock tested positive on a given day. Cell exposure events that occurred within 14 days following a prior cell exposure event were excluded. Cellblock exposure events and events without documented exposures that occurred in the 14 days following a cellblock or cell exposure event were excluded. Facility exposures were stratified by periods of variant predominance (Delta [A]: June 15, 2021 – December 12, 2021; Omicron [B]: December 13, 2021 – May 10, 2022). The associations were estimated using a Cox Proportional Hazard Model stratified by facility and with robust standard errors. The model was adjusted for age, calendar date, race, room and cellblock size, vaccination, and prior infection status of the susceptible person. Boxes indicate estimated hazard ratio (HR) point values and whiskers indicate 95% confidence intervals (Delta Period: n = 19,398 facility events; Omicron Period: n = 14,778 facility events).

| **Supplement Table 13: *Unadjusted estimates of the association between documented exposure and SARS-CoV-2 infection risk among residents incarcerated since beginning*** | | | |
| --- | --- | --- | --- |
| **Type of Facility Exposure** | **Hazard Ratio** | | **95% Confidence Intervals** |
| Delta Period (June 15 to Dec. 12, 2021) | | | |
| No Exposure | - | - | |
| Cellblock Exposure | 5.15 | (3.97-6.69) | |
| Cell Exposures | 33.47 | (23.36-47.95) | |
| Omicron Period (Dec. 13, 2021, to May 10, 2022) | | | |
| No Exposure | - | - | |
| Cellblock Exposure | 5.98 | (4.83-7.40) | |
| Cell Exposures | 12.01 | (9.08-15.89) | |

**Supplement Figure 10: *Effect of prior infection, vaccination, and hybrid immunity on the susceptibility of residents of Connecticut Department of Correction Facilities to SARS-CoV-2 infection between June 15, 2021, and May 10, 2022, by documented SARS-CoV-2 exposure status*** ***(restricted to people incarcerated since beginning of study)***

Legend: Forest plot depicting the association between prior infection, vaccination, and hybrid immunity and the risk of SARS-CoV-2 infection by facility exposure type for residents incarcerated since the beginning of the study (June 15, 2021). Residents were classified as having a cell exposure event (green) the day their cellmate tested positive, having a cellblock exposure event (brown) the day a resident of their cellblock but not cell tested positive, and having an event without documented exposure (navy) if no one in their cellblock tested positive. Cell exposure events that occurred within 14 days following a prior cell exposure event were excluded. Cellblock exposure events and events without documented exposure that occurred in the 14 days following a cellblock or cell exposure event were excluded. Associations were examined using Cox Proportional Hazard Models stratified by cellblock with robust standard errors. Each model was adjusted for age, date of exposure, race, room size, and model (a) was adjusted for vaccination status and model (b) was adjusted for prior infection status. Model (c) was limited to residents with hybrid immunity or residents without a record of prior infection or vaccination. Prior infection was defined as a recorded positive SARS-CoV-2 test ≥90 days before the event and vaccination was defined as the receipt of ≥1 dose before the event. Hybrid immunity was defined as a record of both a prior infection and ≥1 vaccine dose. Boxes indicate estimated hazard ratio (HR) point values and whiskers indicate 95% confidence intervals (Delta: n = 14,535 no exposure events, 4,654 cellblock exposure events, 209 cell exposure events; Omicron: n = 9,808 no exposure events, 4,472 cellblock exposure events, 498 cell exposure events). Ratio of HRs refer to the p-value comparing the HR following cellblock or cell exposure events to the HR following events without documented exposures, estimated using two-sided z-tests. No multiple testing adjustment was performed.

| **Supplement Table 14: *Unadjusted estimates of the effectiveness of prior infection, vaccination, and hybrid immunity on SARS-CoV-2 infection among residents incarcerated since beginning*** | | | | | | | |
| --- | --- | --- | --- | --- | --- | --- | --- |
|  | **Delta Period (June 15 to Dec. 12, 2021)** | | | **Omicron Period (Dec. 13, 2021 - May 10, 2022)** | | | |
| **Type of Facility Exposure** | **Hazard Ratio** | **95% Confidence Intervals** | **Ratio of HR (Pvalue)^a^** | **Hazard Ratio** | **95% Confidence Intervals** | | **Ratio of HR (Pvalue)^a^** |
| Prior SARS-CoV-2 Infection^b^ | | | | | | | |
| No Exposure | 0.20 | (0.11-0.39) | - | 0.31 | | (0.21-0.48) | - |
| Cellblock Exposure | 0.29 | (0.20-0.43) | 0.351 | 0.53 | | (0.43-0.64) | 0.031 |
| Cell Exposures | 0.55 | (0.28-1.05) | 0.036 | 0.69 | | (0.45-0.64) | 0.009 |
| Prior Vaccination^c^ | | | | | | | |
| No Exposure | 0.30 | (0.19-0.48) | - | 0.53 | | (0.36-0.77) | - |
| Cellblock Exposure | 0.32 | (0.23-0.44) | 0.822 | 0.58 | | (0.48-0.70) | 0.663 |
| Cell Exposures | 0.66 | (0.36-1.20) | 0.043 | 0.83 | | (0.55-1.27) | 0.116 |
| Hybrid Immunity^d^ | | | | | | | |
| No Exposure | 0.08 | (0.03-0.21) | - | 0.23 | | (0.14-0.38) | - |
| Cellblock Exposure | 0.11 | (0.06-0.20) | 0.566 | 0.36 | | (0.29-0.46) | 0.036 |
| Cell Exposures | 0.32 | (0.11-0.92) | 0.060 | 0.64 | | (0.39-1.05) | 0.006 |
| ^a^ Hypothesis testing was performed using two-sided z-tests and no multiple comparison adjustment was performed | | | | | | | |
| ^b^ Prior infections were defined as a recorded positive SARS-CoV-2 test at least 90 days prior to the event and vaccination was defined as the receipt of at least one dose prior to the event. | | | | | | | |
| ^c^ Residents were classified as being vaccinated if they had received at least one vaccine dose. | | | | | | | |
| ^d^ Hybrid immunity was defined as a record of both a prior infection and at least one vaccine dose. No infections among cell exposure group. | | | | | | | |

***Restricted to people who tested negative within the five days prior to matching***

In the primary analysis, we include people regardless of recent testing history. This may result in the inclusion of residents who were infected on the day of event. To examine the impact of this, we restricted our sample to residents who tested negative for SARS-CoV-2 within the five days prior to matching.

**Supplement Figure 11: *Association between documented SARS-CoV-2 exposure and SARS-CoV-2 infection risk among residents of Connecticut Department of Correction Facilities between June 15, 2021, and May 10, 2022 (negative in prior five days)***

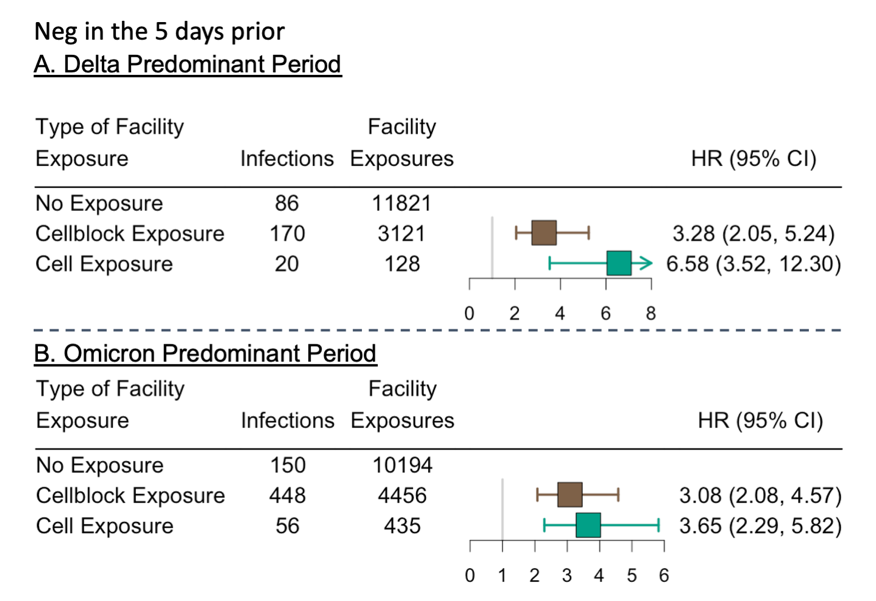
Legend: Forest plot depicting the association between documented close exposure to a SARS-CoV-2 infected resident and the risk of subsequent SARS-CoV-2 infection for residents who tested negative in the five days prior to the facility event. Residents were classified as having a cell exposure event (green) on the day their cellmate tested positive, having a cellblock exposure event (brown) the day a resident of their cellblock but not cell tested positive, and having an event without documented exposures if no one in their cellblock tested positive on a given day. Cell exposure events that occurred within 14 days following a prior cell exposure event were excluded. Cellblock exposure events and events without documented exposures that occurred in the 14 days following a cellblock or cell exposure event were excluded. Facility exposures were stratified by periods of variant predominance (Delta [A]: June 15, 2021 – December 12, 2021; Omicron [B]: December 13, 2021 – May 10, 2022). The associations were estimated using a Cox Proportional Hazard Model stratified by facility and with robust standard errors. The model was adjusted for age, calendar date, race, room and cellblock size, vaccination, and prior infection status of the susceptible person. Boxes indicate estimated hazard ratio (HR) point values and whiskers indicate 95% confidence intervals (Delta Period: n = 15,070 facility events; Omicron Period: n = 15,085 facility events).

| **Supplement Table 15: *Unadjusted estimates of the association between documented exposure and SARS-CoV-2 infection risk among residents who tested negative in the prior 5 days*** | | |
| --- | --- | --- |
| **Type of Facility Exposure** | **Hazard Ratio** | **95% Confidence Intervals** |
| Delta Period (June 15 to Dec. 12, 2021) | | |
| No Exposure | - | - |
| Cellblock Exposure | 6.15 | (4.75-7.98) |
| Cell Exposures | 18.51 | (11.38-30.11) |
| Omicron Period (Dec. 13, 2021 to May 10, 2022) | | |
| No Exposure | - | - |
| Cellblock Exposure | 4.66 | (3.88-5.60) |
| Cell Exposures | 6.09 | (4.49-8.28) |

**Supplement Figure 12: *Effect of prior infection, vaccination, and hybrid immunity on the susceptibility of residents of Connecticut Department of Correction Facilities to SARS-CoV-2 infection between June 15, 2021, and May 10, 2022, by documented SARS-CoV-2 exposure status*** ***(negative in prior five days)***

Legend: Forest plot depicting the association between prior infection, vaccination, and hybrid immunity and the risk of SARS-CoV-2 infection by facility exposure type for residents who tested negative in the five days prior to the facility event. Residents were classified as having a cell exposure event (green) the day their cellmate tested positive, having a cellblock exposure event (brown) the day a resident of their cellblock but not cell tested positive, and having an event without documented exposure (navy) if no one in their cellblock tested positive. Cell exposure events that occurred within 14 days following a prior cell exposure event were excluded. Cellblock exposure events and events without documented exposure that occurred in the 14 days following a cellblock or cell exposure event were excluded. Associations were examined using Cox Proportional Hazard Models stratified by cellblock with robust standard errors. Each model was adjusted for age, date of exposure, race, room size, and model (a) was adjusted for vaccination status and model (b) was adjusted for prior infection status. Model (c) was limited to residents with hybrid immunity or residents without a record of prior infection or vaccination. Prior infection was defined as a recorded positive SARS-CoV-2 test ≥90 days before the event and vaccination was defined as the receipt of ≥1 dose before the event. Hybrid immunity was defined as a record of both a prior infection and ≥1 vaccine dose. Boxes indicate estimated hazard ratio (HR) point values and whiskers indicate 95% confidence intervals (Delta: n = 11,821 no exposure events, 3,121 cellblock exposure events, 128 cell exposure events; Omicron: n = 10,194 no exposure events, 4,456 cellblock exposure events, 435 cell exposure events). Ratio of HRs refer to the p-value comparing the HR following cellblock or cell exposure events to the HR following events without documented exposures, estimated using two-sided z-tests. No multiple testing adjustment was performed.

| **Supplement Table 16: *Unadjusted estimates of the effectiveness of prior infection, vaccination, and hybrid immunity on SARS-CoV-2 infection among residents who tested negative in the prior 5 days*** | | | | | | |
| --- | --- | --- | --- | --- | --- | --- |
|  | **Delta Period (June 15 to Dec. 12, 2021)** | | | **Omicron Period (Dec. 13, 2021 - May 10, 2022)** | | |
| **Type of Facility Exposure** | **Hazard Ratio** | **95% Confidence Intervals** | **Ratio of HR (Pvalue)^a^** | **Hazard Ratio** | **95% Confidence Intervals** | **Ratio of HR (Pvalue)^a^** |
| Prior SARS-CoV-2 Infection^b^ | | | | | | |
| No Exposure | 0.20 | (0.10-0.40) | - | 0.33 | (0.23-0.49) | - |
| Cellblock Exposure | 0.31 | (0.20-0.47) | 0.304 | 0.66 | (0.54-0.81) | 0.002 |
| Cell Exposures | 0.68 | (0.23-2.04) | 0.065 | 0.73 | (0.39-1.36) | 0.033 |
| Prior Vaccination^c^ | | | | | | |
| No Exposure | 0.30 | (0.18-0.48) | - | 0.57 | (0.41-0.79) | - |
| Cellblock Exposure | 0.31 | (0.22-0.45) | 0.844 | 0.8 | (0.66-0.96) | 0.080 |
| Cell Exposures | 0.62 | (0.21-1.85) | 0.228 | 0.73 | (0.43-1.25) | 0.434 |
| Hybrid Immunity^d^ | | | | | | |
| No Exposure | 0.07 | (0.02-0.24) | - | 0.25 | (0.15-0.41) | - |
| Cellblock Exposure | 0.12 | (0.06-0.24) | 0.518 | 0.56 | (0.43-0.73) | 0.005 |
| Cell Exposures | 0.81 | (0.18-3.55) | 0.013 | 0.57 | (0.25-1.30) | 0.102 |
| ^a^ Hypothesis testing was performed using two-sided z-tests and no multiple comparison adjustment was performed | | | | | | |
| ^b^ Prior infections were defined as a recorded positive SARS-CoV-2 test at least 90 days prior to the event and vaccination was defined as the receipt of at least one dose prior to the event. | | | | | | |
| ^c^ Residents were classified as being vaccinated if they had received at least one vaccine dose. | | | | | | |
| ^d^ Hybrid immunity was defined as a record of both a prior infection and at least one vaccine dose. No infections among cell exposure group. | | | | | | |

***Restricted to residents with only one exposure event and censored at time of subsequent events (one SARS-CoV-2 Exposure Analysis)***

We do not require that a person only be exposed to one infected person in the primary analysis. This may result in their being a larger observed effect of documented SARS-CoV-2 exposure than there is in reality. Here, we restrict the sample of events to events with only a single exposure event and censor their time at the time of any subsequent exposure. Residents with cell exposure events were, however, allowed to have a cellblock exposure event and be included in the analysis.

**Supplement Figure 13: *Association between documented SARS-CoV-2 exposure and SARS-CoV-2 infection risk among residents of Connecticut Department of Correction Facilities between June 15, 2021, and May 10, 2022 (one exposure)***

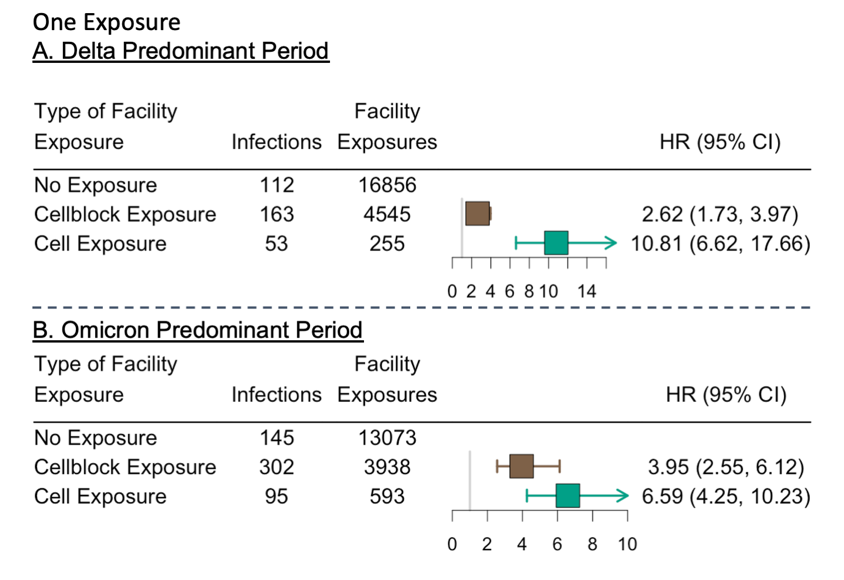
Legend: Forest plot depicting the association between documented close exposure to a SARS-CoV-2 infected resident and the risk of subsequent SARS-CoV-2 infection for residents who experienced only one cell or cellblock event on a given day. Residents were classified as having a cell exposure event (green) on the day their cellmate tested positive, having a cellblock exposure event (brown) the day a resident of their cellblock but not cell tested positive, and having an event without documented exposures if no one in their cellblock tested positive on a given day. Cell exposure events that occurred within 14 days following a prior cell exposure event were excluded. Cellblock exposure events and events without documented exposures that occurred in the 14 days following a cellblock or cell exposure event were excluded. Facility exposures were stratified by periods of variant predominance (Delta [A]: June 15, 2021 – December 12, 2021; Omicron [B]: December 13, 2021 – May 10, 2022). The associations were estimated using a Cox Proportional Hazard Model stratified by facility and with robust standard errors. The model was adjusted for age, calendar date, race, room and cellblock size, vaccination, and prior infection status of the susceptible person. Boxes indicate estimated hazard ratio (HR) point values and whiskers indicate 95% confidence intervals (Delta Period: n = 21,656 facility events; Omicron Period: n = 17,604 facility events).

| **Supplement Table 17: *Unadjusted estimates of the association between documented exposure and SARS-CoV-2 infection risk among residents with one exposure*** | | |
| --- | --- | --- |
| **Type of Facility Exposure** | **Hazard Ratio** | **95% Confidence Intervals** |
| Delta Period (June 15 to Dec. 12, 2021) | | |
| No Exposure | - | - |
| Cellblock Exposure | 4.29 | (3.37-5.46) |
| Cell Exposures | 27.25 | (19.65-37.79) |
| Omicron Period (Dec. 13, 2021 to May 10, 2022) | | |
| No Exposure | - | - |
| Cellblock Exposure | 4.89 | (4.01-5.96) |
| Cell Exposures | 10.40 | (8.03-13.48) |

***Alternative follow-up windows:***

In the primary analysis, we defined follow-up as zero to 14 days after an event. We chose to start the follow-up at time of exposure (the time the index case tested positive) because we believed that the exposure to SARS-CoV-2 from the index case would have likely started a few days prior to when the index case tested positive. We then selected 14 days of follow-up to be in alignment with a recent study by Tan et al. looking at the effect of prior infection and vaccination on infectiousness among residents of California correctional facilities. However, both of these decisions may not reflect the true period of infection risk associated with the index case. Here, we present the results from two sensitivity analyses with alternative follow-up periods.

1. ***Restricted to three to 14 days after matching***
2. ***Restricted to zero to nine days after matching***

***Restricted to three to 14 days after matching***

**Supplement Figure 14: *Association between documented SARS-CoV-2 exposure and SARS-CoV-2 infection risk among residents of Connecticut Department of Correction Facilities between June 15, 2021, and May 10, 2022 (follow up: days 3-14)***

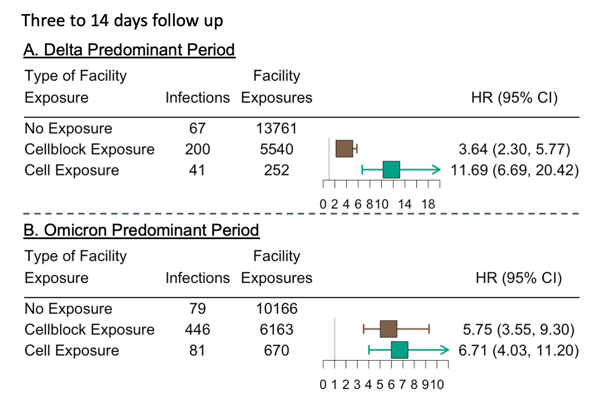
Legend: Forest plot depicting the association between documented close exposure to a SARS-CoV-2 infected resident and the risk of subsequent SARS-CoV-2 infection when follow-up time was defined as 3-14 days after facility exposure event. Residents were classified as having a cell exposure event (green) on the day their cellmate tested positive, having a cellblock exposure event (brown) the day a resident of their cellblock but not cell tested positive, and having an event without documented exposures if no one in their cellblock tested positive on a given day. Cell exposure events that occurred within 14 days following a prior cell exposure event were excluded. Cellblock exposure events and events without documented exposures that occurred in the 14 days following a cellblock or cell exposure event were excluded. Facility exposures were stratified by periods of variant predominance (Delta [A]: June 15, 2021 – December 12, 2021; Omicron [B]: December 13, 2021 – May 10, 2022). The associations were estimated using a Cox Proportional Hazard Model stratified by facility and with robust standard errors. The model was adjusted for age, calendar date, race, room and cellblock size, vaccination, and prior infection status of the susceptible person. Boxes indicate estimated hazard ratio (HR) point values and whiskers indicate 95% confidence intervals (Delta Period: n = 19,553 facility events; Omicron Period: n = 16,999 facility events).

| **Supplement Table 18: *Unadjusted estimates of the association between documented exposure and SARS-CoV-2 infection risk when follow-up time was defined as 3-14 days*** | | |
| --- | --- | --- |
| **Type of Facility Exposure** | **Hazard Ratio** | **95% Confidence Intervals** |
| Delta Period (June 15 to Dec. 12, 2021) | | |
| No Exposure | - | - |
| Cellblock Exposure | 6.74 | (5.11-8.89) |
| Cell Exposures | 32.83 | (22.26-48.43) |
| Omicron Period (Dec. 13, 2021 to May 10, 2022) | | |
| No Exposure | - | - |
| Cellblock Exposure | 8.72 | (6.80-11.17) |
| Cell Exposures | 13.66 | (9.54-18.74) |

**Supplement Figure 15: *Effect of prior infection, vaccination, and hybrid immunity on the susceptibility of residents of Connecticut Department of Correction Facilities to SARS-CoV-2 infection between June 15, 2021, and May 10, 2022, by documented SARS-CoV-2 exposure status*** ***(follow up: days 3-14)***

Legend: Forest plot depicting the association between prior infection, vaccination, and hybrid immunity and the risk of SARS-CoV-2 infection by facility exposure type when follow-up time was defined as 3-14 days after facility exposure event. Residents were classified as having a cell exposure event (green) the day their cellmate tested positive, having a cellblock exposure event (brown) the day a resident of their cellblock but not cell tested positive, and having an event without documented exposure (navy) if no one in their cellblock tested positive. Cell exposure events that occurred within 14 days following a prior cell exposure event were excluded. Cellblock exposure events and events without documented exposure that occurred in the 14 days following a cellblock or cell exposure event were excluded. Associations were examined using Cox Proportional Hazard Models stratified by cellblock with robust standard errors. Each model was adjusted for age, date of exposure, race, room size, and model (a) was adjusted for vaccination status and model (b) was adjusted for prior infection status. Model (c) was limited to residents with hybrid immunity or residents without a record of prior infection or vaccination. Prior infection was defined as a recorded positive SARS-CoV-2 test ≥90 days before the event and vaccination was defined as the receipt of ≥1 dose before the event. Hybrid immunity was defined as a record of both a prior infection and ≥1 vaccine dose. Boxes indicate estimated hazard ratio (HR) point values and whiskers indicate 95% confidence intervals (Delta: n = 13,761 no exposure events, 5,540 cellblock exposure events, 252 cell exposure events; Omicron: n = 10,166 no exposure events, 6,163 cellblock exposure events, 670 cell exposure events). Ratio of HRs refer to the p-value comparing the HR following cellblock or cell exposure events to the HR following events without documented exposures, estimated using two-sided z-tests. No multiple testing adjustment was performed.

| **Supplement Table 19: *Unadjusted estimates of the effectiveness of prior infection, vaccination, and hybrid immunity on SARS-CoV-2 infection when follow-up time was defined as 3-14 days*** | | | | | | | |
| --- | --- | --- | --- | --- | --- | --- | --- |
|  | **Delta Period (June 15 to Dec. 12, 2021)** | | | **Omicron Period (Dec. 13, 2021 - May 10, 2022)** | | | |
| **Type of Facility Exposure** | **Hazard Ratio** | **95% Confidence Intervals** | **Ratio of HR (Pvalue)^a^** | **Hazard Ratio** | | **95% Confidence Intervals** | **Ratio of HR (Pvalue)^a^** |
| Prior SARS-CoV-2 Infection^b^ | | | | | | | |
| No Exposure | 0.22 | (0.10-0.46) | - | 0.35 | | (0.20-0.59) | - |
| Cellblock Exposure | 0.26 | (0.18-0.39) | 0.677 | 0.57 | | (0.47-0.70) | 0.079 |
| Cell Exposures | 0.54 | (0.26-1.12) | 0.091 | 1.05 | | (0.67-1.65) | 0.002 |
| Prior Vaccination^c^ | | | | | | | |
| No Exposure | 0.20 | (0.11-0.37) | - | 0.49 | (0.30-0.77) | | - |
| Cellblock Exposure | 0.31 | (0.23-0.42) | 0.224 | 0.73 | (0.60-0.88) | | 0.114 |
| Cell Exposures | 0.69 | (0.36-1.32) | 0.007 | 1.41 | (0.90-2.21) | | 0.001 |
| Hybrid Immunity^d^ | | | | | | | |
| No Exposure | 0.05 | (0.01-0.24) | - | 0.27 | | (0.15-0.50) | - |
| Cellblock Exposure | 0.08 | (0.03-0.19) | 0.988 | 0.47 | | (0.36-0.61) | 0.106 |
| Cell Exposures | 0.23 | (0.06-0.92) | 0.150 | 1.40 | | (0.76-2.58) | <0.001 |
| ^a^ Hypothesis testing was performed using two-sided z-tests and no multiple comparison adjustment was performed | | | | | | | |
| ^b^ Prior infections were defined as a recorded positive SARS-CoV-2 test at least 90 days prior to the event and vaccination was defined as the receipt of at least one dose prior to the event. | | | | | | | |
| ^c^ Residents were classified as being vaccinated if they had received at least one vaccine dose. | | | | | | | |
| ^d^ Hybrid immunity was defined as a record of both a prior infection and at least one vaccine dose. No infections among cell exposure group. | | | | | | | |

***Restricted to zero to nine days after matching***

**Supplement Figure 16: *Association between documented SARS-CoV-2 exposure and SARS-CoV-2 infection risk among residents of Connecticut Department of Correction Facilities between June 15, 2021, and May 10, 2022 (follow up: days 0-9)***

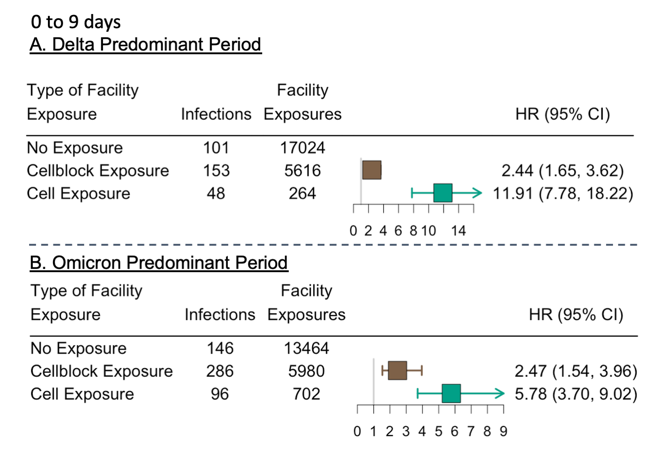
Legend: Forest plot depicting the association between documented close exposure to a SARS-CoV-2 infected resident and the risk of subsequent SARS-CoV-2 infection when follow-up time was defined as 0-9 days after facility exposure event. Residents were classified as having a cell exposure event (green) on the day their cellmate tested positive, having a cellblock exposure event (brown) the day a resident of their cellblock but not cell tested positive, and having an event without documented exposures if no one in their cellblock tested positive on a given day. Cell exposure events that occurred within 14 days following a prior cell exposure event were excluded. Cellblock exposure events and events without documented exposures that occurred in the 14 days following a cellblock or cell exposure event were excluded. Facility exposures were stratified by periods of variant predominance (Delta [A]: June 15, 2021 – December 12, 2021; Omicron [B]: December 13, 2021 – May 10, 2022). The associations were estimated using a Cox Proportional Hazard Model stratified by facility and with robust standard errors. The model was adjusted for age, calendar date, race, room and cellblock size, vaccination, and prior infection status of the susceptible person. Boxes indicate estimated hazard ratio (HR) point values and whiskers indicate 95% confidence intervals (Delta Period: n = 22,904 facility events; Omicron Period: n = 20,146 facility events).

| **Supplement Table 20: *Unadjusted estimates of the association between documented exposure and SARS-CoV-2 infection risk when follow-up time was defined as 0-9 days*** | | | |
| --- | --- | --- | --- |
| **Type of Facility Exposure** | **Hazard Ratio** | | **95% Confidence Intervals** |
| Delta Period (June 15 to Dec. 12, 2021) | | | |
| No Exposure | - | - | |
| Cellblock Exposure | 3.79 | (2.95-4.87) | |
| Cell Exposures | 26.90 | (19.07-37.93) | |
| Omicron Period (Dec. 13, 2021 to May 10, 2022) | | | |
| No Exposure | - | - | |
| Cellblock Exposure | 3.38 | (2.77-4.13) | |
| Cell Exposures | 9.76 | (7.54-12.63) | |

**Supplement Figure 17: *Effect of prior infection, vaccination, and hybrid immunity on the susceptibility of residents of Connecticut Department of Correction Facilities to SARS-CoV-2 infection between June 15, 2021, and May 10, 2022, by documented SARS-CoV-2 exposure status*** ***(follow up: days 0-9)***

| **Supplement Table 21: *Unadjusted estimates of the effectiveness of prior infection, vaccination, and hybrid immunity on SARS-CoV-2 infection when follow-up time was defined as 0-9 days*** | | | | | | |
| --- | --- | --- | --- | --- | --- | --- |
|  | **Delta Period (June 15 to Dec. 12, 2021)** | | | **Omicron Period (Dec. 13, 2021 - May 10, 2022)** | | |
| **Type of Facility Exposure** | **Hazard Ratio** | **95% Confidence Intervals** | **Ratio of HR (Pvalue)^a^** | **Hazard Ratio** | **95% Confidence Intervals** | **Ratio of HR (Pvalue)a** |
| Prior SARS-CoV-2 Infection^b^ | | | | | | |
| No Exposure | 0.16 | (0.08-0.31) | - | 0.32 | (0.22-0.48) | - |
| Cellblock Exposure | 0.23 | (0.15-0.37) | 0.355 | 0.49 | (0.38-0.64) | 0.075 |
| Cell Exposures | 0.52 | (0.26-1.04) | 0.017 | 0.74 | (0.48-1.15) | 0.005 |
| Prior Vaccination^c^ | | | | | | |
| No Exposure | 0.29 | (0.19-0.46) | - | 0.52 | (0.37-0.72) | - |
| Cellblock Exposure | 0.27 | (0.19-0.39) | 0.781 | 0.59 | (0.47-0.74) | 0.522 |
| Cell Exposures | 0.68 | (0.38-1.25) | 0.027 | 0.90 | (0.61-1.35) | 0.034 |
| Hybrid Immunity^d^ | | | | | | |
| No Exposure | 0.06 | (0.02-0.18) | - | 0.21 | (0.13-0.36) | - |
| Cellblock Exposure | 0.11 | (0.06-0.22) | 0.337 | 0.33 | (0.23-0.47) | 0.164 |
| Cell Exposures | 0.28 | (0.09-0.94) | 0.058 | 0.71 | (0.41-1.24) | 0.002 |
| ^a^ Hypothesis testing was performed using two-sided z-tests and no multiple comparison adjustment was performed | | | | | | |
| ^b^ Prior infections were defined as a recorded positive SARS-CoV-2 test at least 90 days prior to the event and vaccination was defined as the receipt of at least one dose prior to the event. | | | | | | |
| ^c^ Residents were classified as being vaccinated if they had received at least one vaccine dose. | | | | | | |
| ^d^ Hybrid immunity was defined as a record of both a prior infection and at least one vaccine dose. No infections among cell exposure group. | | | | | | |

**Directed Acyclic Graph (confounder identification)**

**Supplement Figure 18: *Directed Acyclic Graph (DAG) depicting the relationship between SARS-CoV-2* *exposure, vaccination/prior infection, and SARS-CoV-2 infection***

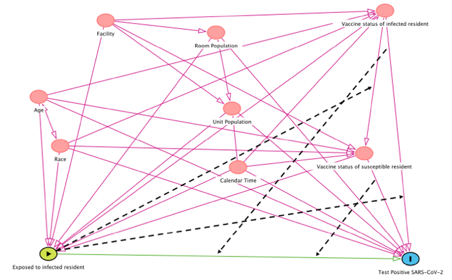
Legend: SARS-CoV-2 exposure is illustrated as the primary exposure of interest (yellow), all pink options are potential confounders (including vaccination and prior infection statuses), and test positive SARS-CoV-2 is the outcome (blue). The black, dashed lines show the potentially modifying nature of SARS-CoV-2 on the association between vaccination/prior infection and SARS-CoV-2.

**References:**

1.	Contact Tracing for COVID-19. *Centers for Disease Control and Prevention* https://www.cdc.gov/coronavirus/2019-ncov/php/contact-tracing/contact-tracing-plan/contact-tracing.html (2022).

2.	Centers for Disease Control and Prevention (CDC). *2020-2021 Nationwide Blood Donor Seroprevalence Survey | Connecticut*. https://covid19serohub.nih.gov/studies/S-1f88deea-873f-4b2a-8ec0-33041516e86c (2021).
